# External validation of plasma CSF1 as a preoperative prognostic marker in patients with resectable intrahepatic cholangiocarcinoma

**DOI:** 10.1101/2024.11.27.24318028

**Authors:** Antonio Akiki, Hanna Jacobsson, Ghada Nouairia, Martin Cornillet, Niklas K. Björkström, Ernesto Sparrelid, Helena Taflin, Hannes Jansson

**Affiliations:** Division of Surgery and Oncology, Department of Clinical Sciences, Intervention and Technology, Karolinska Institutet, Karolinska University Hospital, Stockholm, Sweden; Department of Upper Abdominal Diseases, Karolinska University Hospital, Stockholm, Sweden; Department of Surgery, Institute of Clinical Sciences, Sahlgrenska Academy, University of Gothenburg, Gothenburg, Sweden; Biobank West, Sahlgrenska University Hospital, Gothenburg, Sweden; Unit of Gastroenterology and Rheumatology, Department of Medicine Huddinge (MedH), Karolinska Institutet, Stockholm, Sweden; Center for Infectious Medicine, Department of Medicine Huddinge (MedH), Karolinska Institutet, Stockholm, Sweden; Transplant Center, Region Västra Götaland, Sahlgrenska University Hospital, Gothenburg, Sweden

**Keywords:** intrahepatic cholangiocarcinoma, liver resection, survival, recurrence, prognostic biomarker

## Abstract

**Background & Aims:** Long-term prognosis after resection for intrahepatic cholangiocarcinoma (iCCA) remains poor and the preoperative risk assessment is difficult. A previous single-center study indicated two immune system-related proteins in plasma, colony stimulating factor 1 (CSF1) and TNF-related apoptosis-inducing ligand (TRAIL), as preoperative prognostic factors in iCCA. This study aimed to externally validate CSF1 and TRAIL as prognostic markers for patients with resectable iCCA.

**Methods:** Preoperative plasma CSF1 and TRAIL concentrations (pg/mL) were determined from prospectively collected biobank samples using multiplex immunoanalysis (Proximity Extension Assay), from patients operated for iCCA at two tertiary referral centers, Karolinska (2010-2020) and Sahlgrenska (2019-2023) university hospitals. The primary outcome was overall survival (OS), analyzed by Kaplan-Meier method and Cox regression. Secondary outcome was disease-free survival (DFS).

**Results:** Sixty-one patients with resection for iCCA were included. CSF1 above median was associated with lymph node metastasis (P=0.03). CSF1 was associated with both OS (hazard ratio [HR] 1.03, 95% confidence interval [CI] 1.01-1.05) and DFS (HR 1.02, 95% CI 1.00-1.04). The median OS was eight months for patients with CSF1 values in the upper quartile (≥158 pg/mL), compared to an overall median OS of 47 months. While TRAIL was not significantly associated with OS (P=0.22), values in the lower quartile (≤256 pg/mL) were associated with short DFS (P<0.01). Multivariable analyses confirmed the independent prognostic significance of CSF1. The C-index of CSF1 for OS was 0.70, with excellent calibration for three- and five-year OS.

**Conclusion:** Plasma CSF1 was validated as a novel independent, well-calibrated preoperative predictor of poor survival in resectable iCCA, which could assist the preoperative risk assessment. Low plasma TRAIL was associated with early recurrence.

**Impact and implications:** Patients with intrahepatic cholangiocarcinoma (iCCA) suffer a high risk of recurrence within the first years after curative intent surgery, limiting long-term survival. To identify patients where liver surgery has a low potential for cure, prognostic markers are warranted. In the present study, preoperative plasma CSF1, an immune system-related protein, was validated as a novel, well-calibrated predictor for long-term survival after hepatic resection. Plasma CSF1 could assist the preoperative risk assessment in iCCA.

## INTRODUCTION

The long-term prognosis after curative intent resection for intrahepatic cholangiocarcinoma (iCCA) is poor, with a median survival of less than five years [1]. Hepatic resection for iCCA carries a considerable risk of postoperative morbidity [2,3]. However, the preoperative risk assessment for patients with iCCA remains difficult and neoadjuvant protocols have not yet been established as a standard of care [4]. An improved preoperative assessment of prognosis could aid in choosing a more individualized treatment strategy for patients with iCCA, e.g. identifying candidates for up-front surgery or neoadjuvant therapy.

An inflammation response plays a central role in the development and progression of cancer. Chronic inflammation is a risk factor in developing cancer, but also plays a key role in cancer progression through the recruitment and infiltration of immune cells in the tumor microenvironment and the dysregulation of the immune response, facilitating tumor growth [5–7]. Preoperative plasma concentrations of general inflammatory parameters (c-reactive protein [CRP], albumin) have been associated with survival in cancer, and validated as negative prognostic factors in patients with biliary tract cancer, but are not specific to iCCA [8–10]. A previous single-center biomarker discovery study indicated that colony stimulating factor 1 (CSF1), a cytokine that controls the production, differentiation, and function of macrophages, and also tumor necrosis factor-related apoptosis-inducing ligand (TRAIL), a cytokine that induces apoptosis through binding to death receptors, have potential as preoperative prognostic factors for patients with iCCA [11].

This study aimed to validate the immunological plasma markers CSF1 and TRAIL as preoperative prognostic factors for patients with resectable iCCA, in collaboration between Sweden’s two largest regional referral centers.

## METHODS

### Study design

Patients scheduled for resection due to iCCA at two tertiary referral centers, Karolinska (Stockholm, Sweden) and Sahlgrenska (Gothenburg, Sweden) university hospitals, were prospectively included in institutional biobanks. All patients with available samples, not previously analyzed, were included in this validation study. In the preceding biliary tract cancer biomarker discovery study, patients operated for iCCA at Karolinska university hospital between January 2010 and January 2017 were included by randomized selection [11]. All iCCA patients not analyzed in the previous study were included in the validation cohort. Furthermore, all patients with available biobank samples at Karolinska university hospital from February 2017 to 2020, and at Sahlgrenska university from 2019 to 2023 were included. The study was reported in accordance with REMARK guidelines [12]. The REMARK checklist is presented in Supplemental Table 1.

### Sample size analysis

Sample size was calculated prior to study initiation, according to relative risk data for the iCCA group derived from the biomarker discovery study [11]. With an estimated median survival time of two years, a median follow-up time of four years and a yearly censoring rate of five percent, a sample size of n=51 was estimated as necessary to detect a survival difference according to CSF1 concentrations above or below median (alpha = 0.05, beta = 0.20, hazard ratio [HR] 2.39) [13].

### Patient inclusion

Characteristics of the biobank cohort and patient inclusion is presented in **Figure 1**. The biobank cohort consisted of 69 patients assessed as candidates for curative intent resection of iCCA. Sixty-two patients underwent resection, while seven were found to have an unresectable tumor upon surgical exploration. One patient with resection in the biobank had a final histopathological diagnosis of hepatocellular cancer and was excluded. The primary validation cohort included 61 resected patients. Sensitivity analyses were performed including patients in the unresected group. All patients provided written consent to store their plasma samples in the biobank. The study was approved by the Swedish Ethical Review Authority/Regional Ethical Review Board of Stockholm with reference numbers 2022-03763-01, 2020-02702 and 2013/188-31.

**Figure 1.**
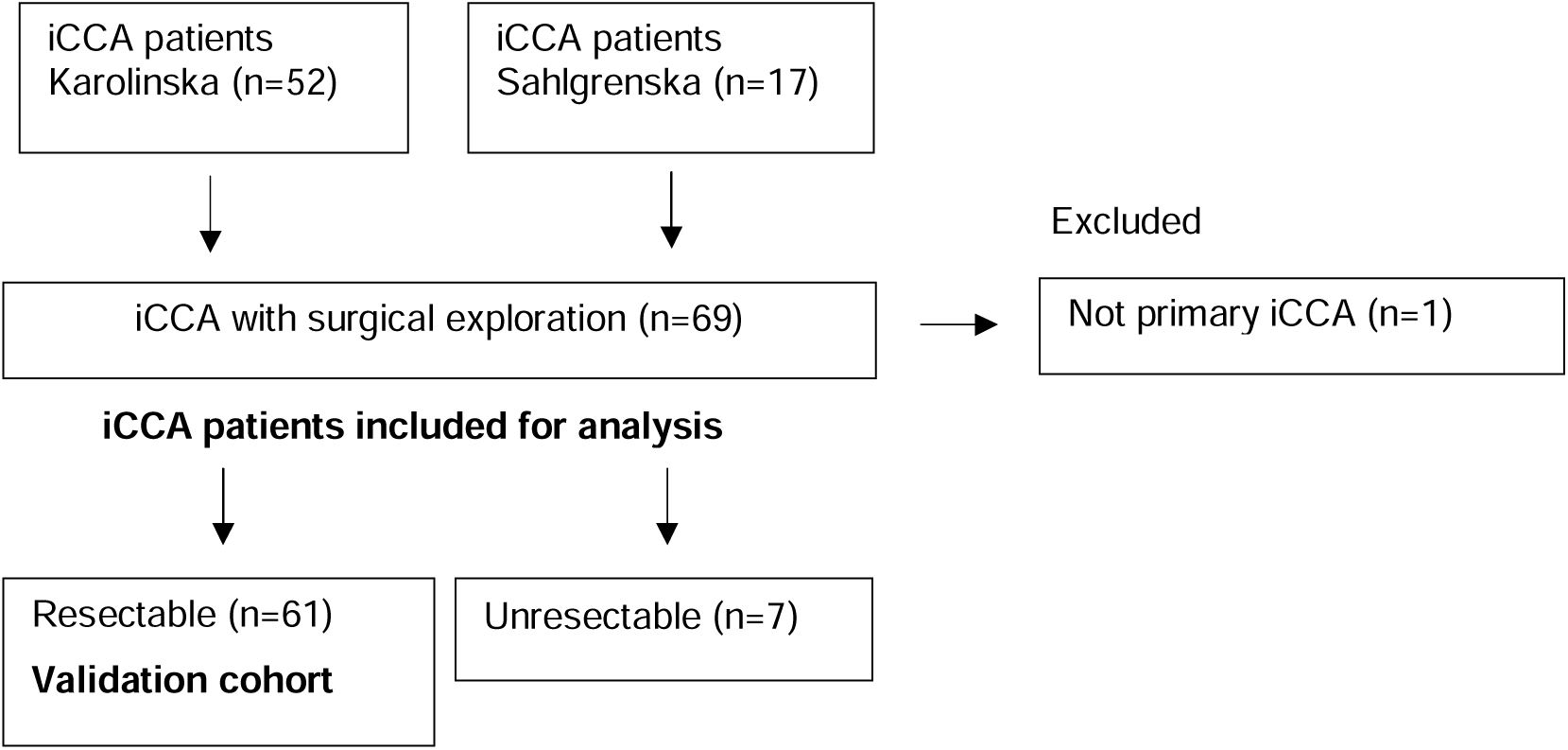
Patient inclusion. iCCA, intrahepatic cholangiocarcinoma.

### Outcome variables and clinical data

The primary outcome was overall survival (OS), which was calculated from the date of surgery to the date of death or last follow-up. The secondary outcome was disease-free survival (DFS), calculated from the date of surgery to the event of either recurrence (according to clinical/radiological follow up) or death [14]. Data were collected retrospectively from quality registries and the electronic health records. Clinical variables collected included: sex, age, body mass index (BMI), American Society of Anesthesiologists’ (ASA) classification for preoperative physical status, comorbidity (diabetes, primary sclerosing cholangitis [PSC], inflammatory bowel disease [IBD]), CRP, albumin, carbohydrate antigen 19-9 (CA19-9) and histopathological AJCC/TNM classification [15]. Electronic health records were linked to the Swedish national population registry for data on vital status. Last date of follow up was 4 March 2024.

### Sample preparation and protein quantification

Plasma samples were drawn preoperatively the day of surgery, centrifuged, frozen, and stored in biobank at -80 degrees Celsius. Quantitative Proximity Extension Assay (PEA) was used to determine preoperative CSF1 and TRAIL concentrations and was performed by the Affinity Proteomics Stockholm core facility (SciLifeLab, Stockholm, Sweden), blinded to all clinical and outcome data, using the Olink® Target 48 Cytokine panel (Olink Proteomics AB, Uppsala, Sweden). Twenty-five microliters of plasma were added to each well of two 48 well plates (Thermo Fisher Scientific Inc. Mass. USA). PEA utilizes antibody pairs that target their intended proteins. The paired antibodies are labelled with specific and complementary oligonucleotides allowing quantification of protein concentrations by polymerase chain reaction [16,17]. None of the analyzed samples failed quality control and all were above the limit of detection. Ten of the plasma samples were randomly chosen and analyzed on both plates to check for variances in detected protein concentrations and the observed variance was less than 25% which was deemed acceptable.

### Statistical analysis

Statistical analysis was performed in R (RStudio v 2024.04.0+735, R v 4.3.2 [2023-10-31]) and SPSS Statistics v 28 (IBM, New York, USA). R packages used were: survival (v 3.6-4), rms (v 6.7-0), survminer (v 0.4.9), ggplot2 (v 3.5.1) and dplyr (v 1.1.4). Uni- and multivariable analyses were performed using Cox regression. Scaled Schoenfeld residuals were used to test the proportional hazards assumption. Survival times were visualized using Kaplan-Meier curves and survival curves compared by log-rank test. Concordance indices (C-index) were used to evaluate the predictive value of independent variables, where a C-index of 0.5 would indicate no predictive ability and an index of 1.0 a perfect predictive ability. Internal validation of prognostic models was performed with bootstrap validation (500 resamples) to account for overfitting [18]. Predictive performance was assessed with calibration curves. Differences in baseline characteristics were analyzed using Chi-Square test or Fisher’s exact test as appropriate for binary variables, and with Mann-Whitney-Wilcoxon test for continuous variables. A two-sided P-value <0.05 was considered statistically significant.

## RESULTS

### Baseline characteristics

Sixty-one patients, 34 men and 27 women, were included in the validation cohort. The median age was 62 years (IQR 62-74 years). The median CSF1 and TRAIL concentrations (IQR) were 139.32 (130.05-154.49) and 318.12 (262.81-372.62) pg/mL, respectively. A comparison of baseline characteristics according to CSF1 and TRAIL concentrations (above/below median), are presented in **Table 1**. There was a significantly higher occurrence of lymph node metastasis (P=0.03) and higher CRP levels (P<0.01) in patients with CSF1 above median. The plasma TRAIL level was associated with the albumin concentration (P=0.02), and negatively associated with CA19-9 (P=0.04). For the remaining variables, no statistically significant differences were observed between the groups. No significant differences in baseline characteristics or in CSF1 (P=0.72) and TRAIL concentrations (P=0.73) where seen when comparing the resectable and unresectable groups (**Supplemental Table 2**).

**Table 1.** Comparison of baseline characteristics according to plasma CSF1 and TRAIL concentration (above/below median)

|  | Missing data (n) | CSF1 (pg/mL) below median (n=30) | CSF1 (pg/mL) above median (n=31) | P-value | TRAIL (pg/mL) below median (n=30) | TRAIL (pg/mL) above median (n=31) | P-value |
| --- | --- | --- | --- | --- | --- | --- | --- |
| Age, md (IQR) | 0 | 68 (63.25-72.75) | 71 (62-75) | 0.97<sup>§</sup> | 68 (63.5-71.75) | 71 (61.5-75.0) | 0.39<sup>§</sup> |
| Sex (male), n (%) | 0 | 18 (60) | 16 (52) | 0.69 | 17 (57) | 17 (57) | 1.00 |
| BMI, md (IQR) | 0 | 25,5 (21.3-29.6) | 27.8 (24.3-30.7) | 0.09<sup>§</sup> | 25.5 (23.6-29.2) | 27.8 (23.1-30.7) | 0.19<sup>§</sup> |
| ASA ≥3, n (%) | 0 | 7 (23) | 13 (42) | 0.20 | 11 (37) | 9 (29) | 0.72 |
| Diabetes, n (%) | 0 | 7 (23) | 9 (30) | 1.00<sup>#</sup> | 8 (26) | 8 (27) | 1.00<sup>#</sup> |
| PSC, n (%) | 0 | 0 (0) | 2 (7) | 0.57<sup>#</sup> | 2 (6) | 0 (0) | 0.53<sup>#</sup> |
| IBD, n (%) | 0 | 3 (10) | 3 (10) | 1.00<sup>#</sup> | 3 (10) | 3 (10) | 1.00<sup>#</sup> |
| Albumin g/L, md (IQR) | n=11 | 38 (36-40) | 38 (33.5-39.5) | 0.25<sup>§</sup> | 36.5 (33.75-39) | 39 (38-40) | **0.02<sup>§</sup>** |
| CRP mg/L, md (IQR) | n=9 | 2 (1-3) | 5.5 (3-26.25) | **<0.01<sup>§</sup>** | 3.5 (2-16) | 3 (1-4.25) | 0.20<sup>§</sup> |
| CA19-9 kU/L, md (IQR) | n=25 | 21 (15-159.5) | 40 (12-1230) | 0.20<sup>§</sup> | 41 (30-584) | 15 (7.3-459) | **0.04<sup>§</sup>** |
| T ≥3, n (%) | 0 | 10 (33) | 14 (45) | 0.49 | 14 (47) | 10 (32) | 0.37 |
| N1, n (%) | 0 | 2 (7) | 10 (32) | **0.03** | 7 (23) | 5 (16) | 0.70 |
| Adjuvant therapy, n (%) | n=4 | 12 (39) | 17 (57) | 0.23 | 13 (42) | 16 (53) | 0.51 |

**Abbreviations:** BMI, body mass index. ASA≥3, American Society of Anesthesiologists physical status classification 3-4. PSC, primary sclerosing cholangitis. IBD, inflammatory bowel disease. CRP, c-reactive protein. CA19-9, carbohydrate antigen 19-9. T≥3, tumor extension 3-4. N1, lymph node metastasis. P-values <0.05 in bold. § Mann-Whitney-Wilcoxon test. # Fisher's exact test.

### Overall survival and disease-free survival

The median follow-up time was 68 months (95% CI 47-90 months). During follow-up, 29 deaths and 29 confirmed recurrences were observed. The median OS time was 47 months (95% CI 11-84 months), with a 3-year OS of 56% (95% CI 44-71 months) and 5-year OS 49% (95% CI 37-65 months). The median DFS time was 17 months (95% CI 4-29 months), with a 3-year DFS of 41% (95% CI 29-56 months) and 5-year DFS 38% (95% CI 26-54 months). Three patients were lost to follow up postoperatively regarding recurrence data and were coded as recurrence-free the day of discharge from the hospital.

Cox regression analyses for OS and DFS are presented in **Table 2**. In the univariable Cox regression higher plasma concentrations of CSF1 showed a significant negative association with OS (P=0.001), with a risk increase in mortality of 3% per unit of concentration (pg/mL) and a C-index of 0.71. Plasma TRAIL concentrations were not significantly associated with OS (P=0.22).

**Table 2.**
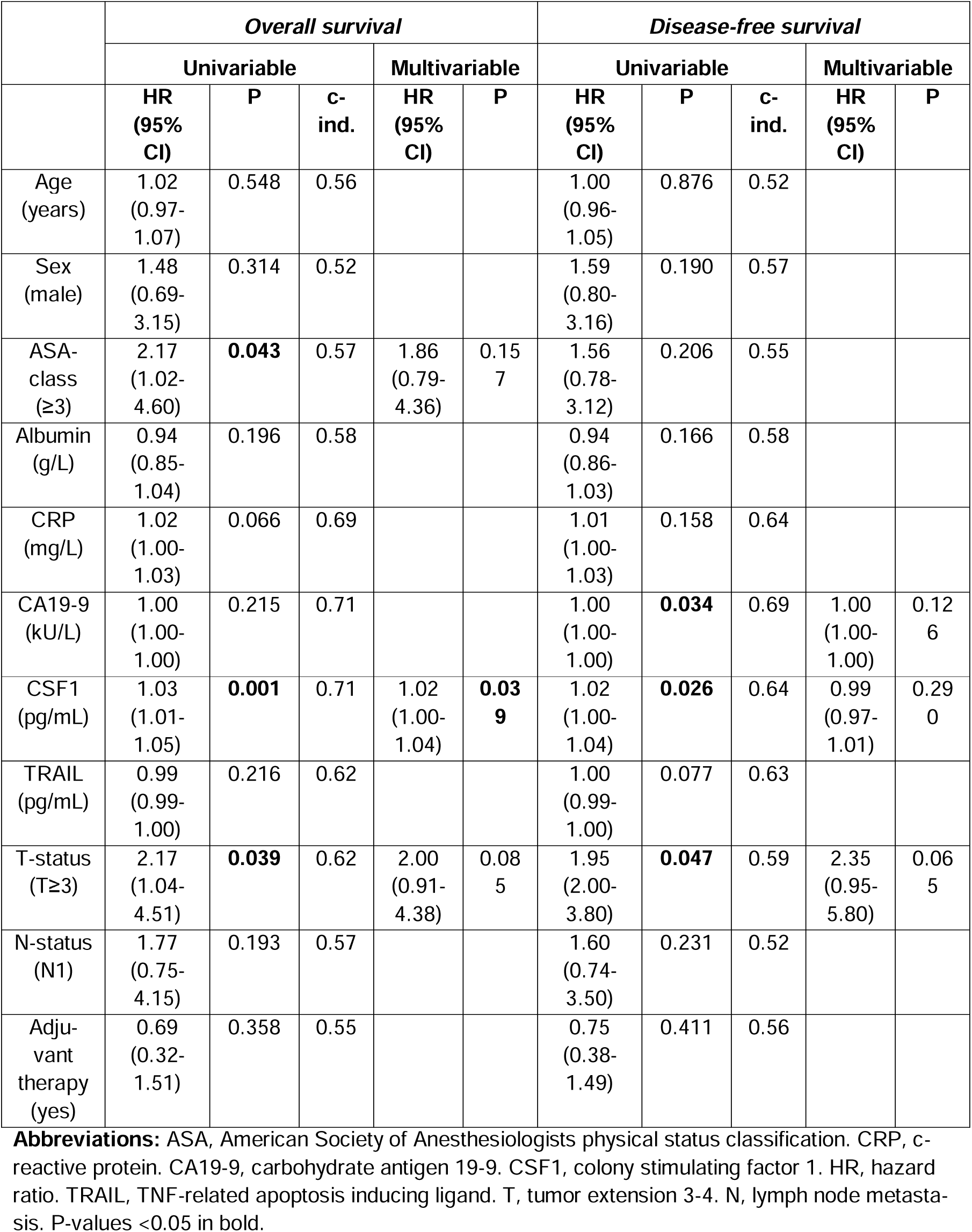
Uni- and multivariable Cox regression analyses for overall survival (OS) and disease-free survival (DFS)

The preoperative ASA-class (P=0.043) and postoperative histopathological T-status (P=0.039) were negatively associated with OS, with C-indices of 0.57 and 0.62 respectively. On multivariable analysis, a model including CSF1, ASA and T-status had a cumulative C-index of 0.71 (P<0.001), with only CSF1 remaining independently associated with OS (P=0.039, HR 1.02, 95% CI 1.00-1.04).

Elevated CSF1 levels were negatively associated with DFS (P=0.026, HR 1.02, 95% CI 1.00-1.04), with a C-index of 0.64. TRAIL as a continuous variable was not significantly associated with DFS (P=0.077).

Furthermore, the histopathological T-status and preoperative CA19-9 were associated with DFS. On multivariable analysis including CSF1, CA19-9 and T-status, CSF1 was not independently associated with DFS (P=0.29). The combined model had a cumulative C-index of 0.67 (P=0.03) for DFS.

To account for overfitting, bootstrap validation was performed. Plasma CSF1 had corrected C-indices of 0.70 for OS (0.71 uncorrected), and 0.64 for DFS (0.64 uncorrected).

OS was also analyzed according to CSF1 and TRAIL concentrations stratified by quartiles (**Figure 2**). CSF1 levels by quartiles were significantly associated with OS (P<0.001). Patients with CSF1 concentrations in the upper quartile (Q4, blue line) had a median OS of 8 months (95% CI 0-19 months), compared to an overall OS of 47 months for the whole population. For patients with CSF1 levels in the lower quartile (Q1, green line) median OS was not reached. Plasma TRAIL quartiles showed an insignificant association with OS (P=0.07).

**Figure 2.**
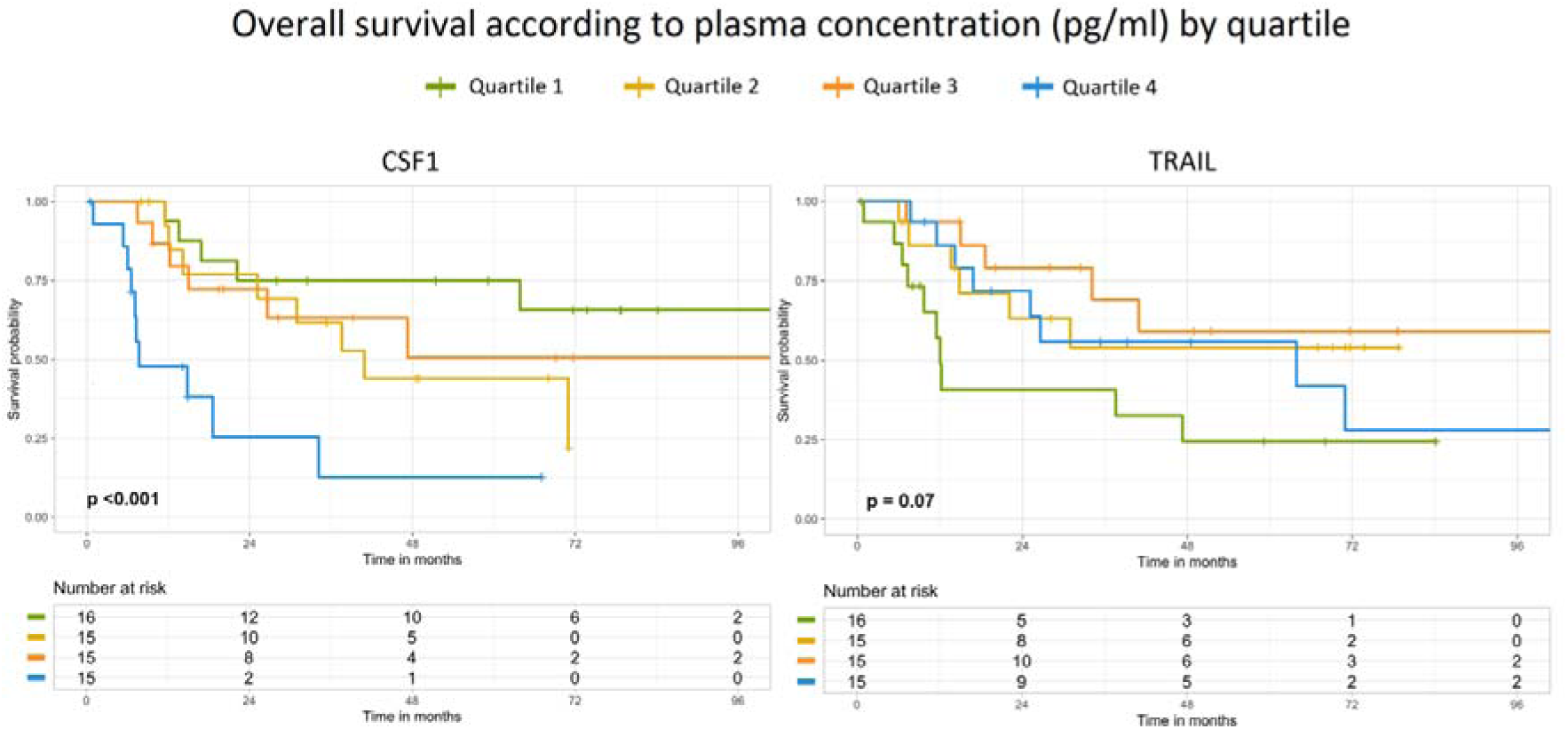
Overall survival according to CSF1 and TRAIL concentrations (pg/mL) stratified by quartiles (Q1 green; Q2 yellow; Q3 orange; Q4 blue lines). CSF1, colony stimulating factor 1. TRAIL, TNF-related apoptosis inducing ligand. P-values by log-rank test.

Plasma CSF1 and TRAIL concentrations stratified according to quartiles were significantly associated with DFS **(Figure 3)**. For CSF1, median DFS in the upper quartile (Q4, blue line) was seven months (95% CI 5-8 months), compared to a median DFS of 62 months for patients in the lower quartile (Q1, green line). Patients with TRAIL concentrations in the lower quartile (Q1, green line) had a median DFS of 9 months (95% CI 6-11 months), significantly shorter compared to patients in the second to fourth quartile (P<0.01).

**Figure 3.**
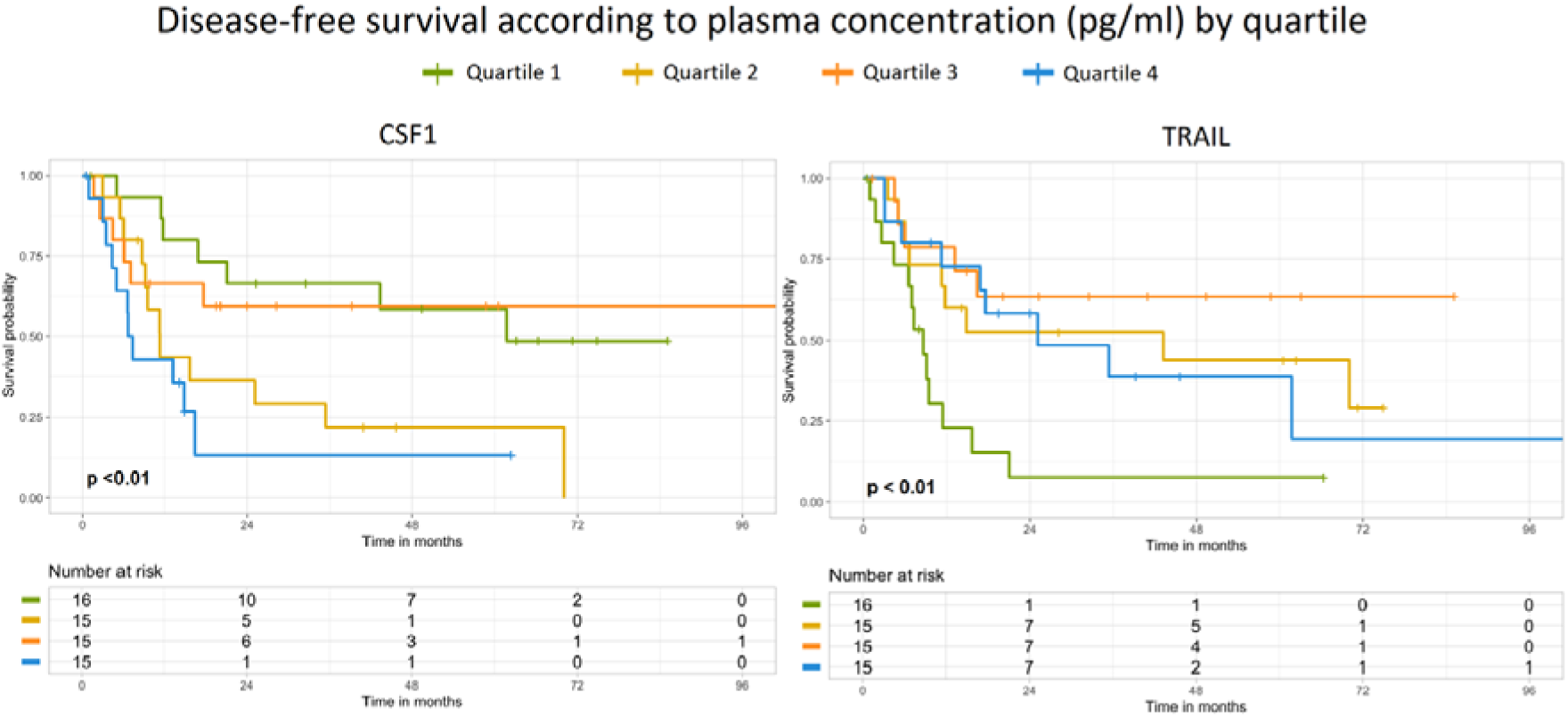
Disease-free survival according to CSF1 and TRAIL concentrations (pg/mL) stratified by quartiles (Q1 green; Q2 yellow; Q3 orange; Q4 blue lines). CSF1, colony stimulating factor 1. TRAIL, TNF-related apoptosis inducing ligand. P-values by log-rank test.

### Sensitivity analyses

The validation cohort consisted of patients undergoing resection. In a sensitivity analysis including all operated patients from the biobank cohort (n=68; resectable n=61, unresectable n=7) CSF1 remained significantly associated with OS (P=0.001) with a HR of 1.03 (95% CI 1.01-1.05) and a C-index of 0.70, while TRAIL remained without significant association with OS (P=0.193, HR 1.00, 95% CI 0.99-1.00).

In the primary analysis, patients with tumors classified as Nx i.e. no lymph node dissection performed (n=27, 44%), were analyzed together with patients staged with N0-status (no lymph node metastasis). A sensitivity analysis was performed excluding patients staged as Nx. Lymph node metastasis remained significantly more prevalent among patients with CSF1 levels above the median (P=0.01), while N-status remained non-significantly associated with OS (P=0.10) and DFS (P=0.20).

The predictive ability of CSF1 for OS and DFS was compared with that of Glasgow prognostic score (GPS), a scoring tool incorporating CRP and albumin [18], and that of preoperative CA19-9 (above the upper normal reference limit 35 kU/L) [20] **(Supplemental Table 3)**. An elevated GPS (i.e. albumin<35g/L and/or CRP>10mg/L) and elevated CA19-9 (≥35 kU/L) were both negatively associated with OS (P=0.02, C-index=0.66 and P=0.003, C-index=0.69, respectively). While an elevated CA19-9 (≥35 kU/L) was negatively associated with DFS (p=0.001, C-index=0.69), no significant association with DFS was seen for GPS (P=0.07).

The combination of CSF1 and GPS (P=0.01) had a C-index of 0.75 (0.73 corrected) for OS, while CSF1 and CA19-9 (≥35 kU/L) (P=0.03) had a C-index of 0.70 (0.67 corrected) for OS. Combining CSF1, GPS and CA19-9 (≥35 kU/L) (P=0.02) yielded a C-index for OS of 0.74 (0.69 corrected). Elevated GPS was significantly more prevalent in patients with CSF1 concentrations above the median, suggesting a correlation between the variables (P<0.01).

As patients with CSF1 concentrations in the upper quartile (Q4, **Figure 2**) had markedly worse OS, the predictive value of using the upper quartile limit (>158 pg/mL) as a binary cutoff was analyzed (P<0.001), with a C-index of 0.65 (0.65 corrected) for OS **(Supplemental Table 3)**.

Further potential prognostic models were evaluated, and are presented in **Supplemental Table 4**. A preoperative model combining CSF1, GPS, CA19-9 (≥35 kU/L) and ASA-class (≥3) to assess OS (P=0.02) achieved a C-index of 0.73 (0.65 corrected). Missing data, primarily for preoperative CA19-9, resulted in a higher degree of overfitting, as observed in the bootstrap validated corrected C-indices <0.70.

Omitting CA19-9, the combination of CSF1, GPS and ASA as a preoperative model for OS (P<0.01) yielded a C-index of 0.76 (0.72 corrected). Adding histopathological T-status for a comprehensive prognostic model for OS (P<0.01) yielded a C-index of 0.81 (0.76 corrected).

### Calibration

The predictive performance of preoperative CSF1 for one-, three- and five-year OS was assessed with calibration curves **(Supplemental Figure 1)** [18]. Blue lines illustrate the bootstrap corrected prognostic model and black lines the uncorrected model. With CSF1 as a prognostic factor, the actual survival at the one-year mark was lower than predicted. Plasma CSF1 was well calibrated as a prognostic model for three- and five-year survival, with highly accurate predictions across all probability ranges for five-year survival.

Calibration was assessed for a model combining CSF1 and GPS to predict one-, three- and five-year survival **(Supplemental Figure 2)**. For one-year survival the model was optimistic, overestimating survival. For three- and five-year survival the model underestimated actual survival predicted below 60%, and overestimated survival predicted above 60%.

Calibration analyses for a preoperative prognostic model including CSF1, ASA and GPS and a comprehensive prognostic model including CSF1, ASA, GPS and T-status are presented in **Supplemental Figure 3** and **Supplemental Figure 4**, respectively. Both models were poorly calibrated for one-, three- and five-year survival.

A model combining CSF1 and ASA performed better than CSF1 alone in predicting one-year survival (**Supplemental Figure 5**), while underestimating longer term survival predicted below approximately 40%.

## DISCUSSION

The long-term prognosis for patients undergoing curative intent resection for iCCA is poor, with a 5-year OS of less than 50%. Preoperative risk assessment remains difficult, with a limited sensitivity for current radiological staging modalities [4].

In this validation cohort of patients resected for histopathologically verified iCCA at Sweden’s two largest tertiary referral centers, higher preoperative levels of the immunological plasma marker CSF1 were associated with earlier recurrence and shorter survival time, with a 3% increase in mortality per unit of concentration. Patients with CSF1 levels in the upper quartile had a median OS of only eight months after resection, compared to an overall median OS time of 47 months. In multivariable analysis CSF1 was an independent prognostic factor. The predictive ability of CSF1 was good, and the marker was well-calibrated to assess long-term survival.

TRAIL levels were not significantly associated with OS or DFS in univariable Cox regression analysis. In analysis of DFS according to TRAIL concentration by quartiles, a significant association was seen for patients with the lowest preoperative TRAIL levels in plasma. In the lower TRAIL quartile, 15 out of 16 patients suffered cancer recurrence or death within two years of resection. The survival curves for the second to fourth quartiles crossed multiple times and were of low prognostic value. This suggests that while higher TRAIL levels do not effectively differentiate DFS risk, low TRAIL levels are associated with early recurrence.

Lymph node metastasis was significantly more prevalent in patients with CSF1 concentrations above the median, indicating a direct link between disease severity and CSF1 levels. That lymph node metastasis on its own was not a significant prognostic factor in this analysis could be a consequence of the large proportion of patients operated without lymph node dissection in this cohort (Nx classification, i.e. unknown lymph node status). When analyzing only patients with known lymph node status, the sample size and the statistical power to detect an association between N1 status and OS or DFS was further restricted. While CSF1 levels were associated with N1-status and the preoperative CRP level, TRAIL was associated with preoperative albumin and CA19-9.

Glasgow prognostic score, a composite score based on CRP and albumin, has previously been validated as an independent preoperative prognostic factor for patients with biliary tract cancer [8]. GPS was significantly associated with OS also in this cohort of patients with resectable iCCA. While GPS and CSF1 levels were found to be associated, the combination of GPS and CSF1 provided improved predictive ability for OS, albeit with less well-calibrated predictions.

Our results showed that CSF1, as a continuous variable, was well-calibrated as a prognostic model for three- and five-year survival, though its accuracy was lower for one-year survival. The predictive ability and precision decreased when CSF1 was combined with additional variables (**Supplemental Figures 2-4**), with the exception of an improved predictive ability for one-year survival when CSF1 was combined with preoperative ASA-class (**Supplemental Figure 5**). This underscores how preoperative physical status significantly impacts short-term survival in iCCA patients, while CSF1 is an accurate predictor of longer-term outcomes.

Both CSF1 and TRAIL have been studied as disease-specific markers of macrophage and lymphocyte activity in tumor and peritumoral tissue in iCCA. High expression of CSF1 is associated with increased tumor associated macrophage (TAM) infiltration to promote tumor growth, angiogenesis and metastasis [21]. A previous pre-clinical in vivo study explored the effect of CSF1 receptor (CSF1-R) inhibition on recruitment of tumor associated macrophages and CCA growth. The study found that while CSF1-R inhibition reduced TAM recruitment, it did not slow tumor growth due to a compensatory infiltration of granulocyte-like myeloid suppressor cells [22]. A second study showed that combination therapy with a CSF1-R inhibitor and ALOX-5 (an enzyme involved in lipid signaling) reduced both macrophage infiltration to the tumor microenvironment and tumor size in a mouse xenograft model [23].

TRAIL induces apoptosis through binding to death receptors DR4 and DR5 with a high specificity to tumor cells, and has been studied as a potential targeted oncological therapy. However, studies exploring TRAIL receptor agonists have yielded mixed results. While TRAIL has been shown to induce apoptosis in various types of cancer, several cholangiocarcinoma cell lines are resistant to TRAIL signaling [24,25]. A recent study indicated a dual role for TRAIL in the CCA tumor microenvironment, in the sense that noncanonical TRAIL signaling promotes tumor growth by increasing the accumulation of myeloid suppressor cells, and that lower tissue TRAIL-receptor expression was associated with improved survival [26]. The prognostic impact of plasma CSF1 and the association of low TRAIL with short DFS seen in this study, motivates further investigation of the potential association of plasma CSF1 and TRAIL with the iCCA tumor microenvironment.

Through a bi-institutional biobank cohort, this study enabled the validation of CSF1 as an independent and well-calibrated preoperative prognostic factor for patients with iCCA. The long median follow-up time and complete survival data were important strengths of the study.

There were also a number of limitations to this study. While the validation sample size was estimated according to data from the preceding biomarker discovery study, the cohort was relatively small, and the sample size calculated according to relative risk data for CSF1. Therefore, for the negative finding for TRAIL a type II error cannot be excluded, and further study is necessary. Another limitation is that while patients were prospectively included in biobanks, clinical follow-up data were retrospectively collected from charts and registries, with incomplete registration including loss to follow-up for recurrence status. Furthermore, with only preoperative plasma samples available, temporal dynamics of protein expression after resection could not be evaluated. Lastly, further study is motivated to assess the predictive ability of CSF1 in different geographical regions with a higher prevalence of risk factors such as viral hepatitis, endemic parasitic infections and hepatolithiasis [27,28].

While hepatic resection remains the only established curative treatment for patients with iCCA [4], surgery can carry a significant risk of postoperative morbidity and has a high rate of cancer recurrence and mortality within the first three to five years post-operatively [1]. Plasma CSF1 has the potential to aid in preoperative identification of high-risk patients less likely to benefit long-term from up front surgery, support individualized risk-benefit analyses and guide patient inclusion for therapeutic trials, including neoadjuvant or targeted approaches. To further assess plasma CSF1 as a preoperative prognostic marker for patients with resectable iCCA, validation in prospective cohorts is motivated.

In conclusion, plasma CSF1 was validated as a novel, independent and well-calibrated preoperative prognostic marker, which could assist a preoperative risk stratification for patients with resectable iCCA. While not significantly associated with survival, low TRAIL was associated with DFS and should be further investigated as a marker of early recurrence.

## Data Availability

All data produced in the present study are available upon reasonable request to the authors

## Abbreviations

ASA: American Society of Anesthesiologists’ physical status classification
BMI: body mass index
CA19-9: carbohydrate antigen 19-9
C-index: concordance index
CI: confidence interval
CCA: cholangiocarcinoma
CRP: c-reactive protein
CSF1: colony-stimulating factor 1
DFS: disease-free survival
GPS: Glasgow prognostic score
HR: hazard ratio
IBD: inflammatory bowel disease
iCCA: intrahepatic cholangiocarcinoma
N1: lymph node metastasis
OS: overall survival
PEA: proximity extension assay
pg/mL: picogram per millilitre
PSC: primary sclerosing cholangitis
TNM: TNM Classification of Malignant Tumors 7^th^ Ed.
TRAIL: TNF-related apoptosis inducing ligand

## Acknowledgements

The authors would like to thank SciLifeLab Affinity Proteomics Stockholm; Biobank West; Biobank and Study Support at Karolinska University Hospital – Stockholm Medical Biobank; and Jenny Höök and Christina Klarås Nilsson (Karolinska University Hospital) for their support.

---

**Supplemental Figure 1.**
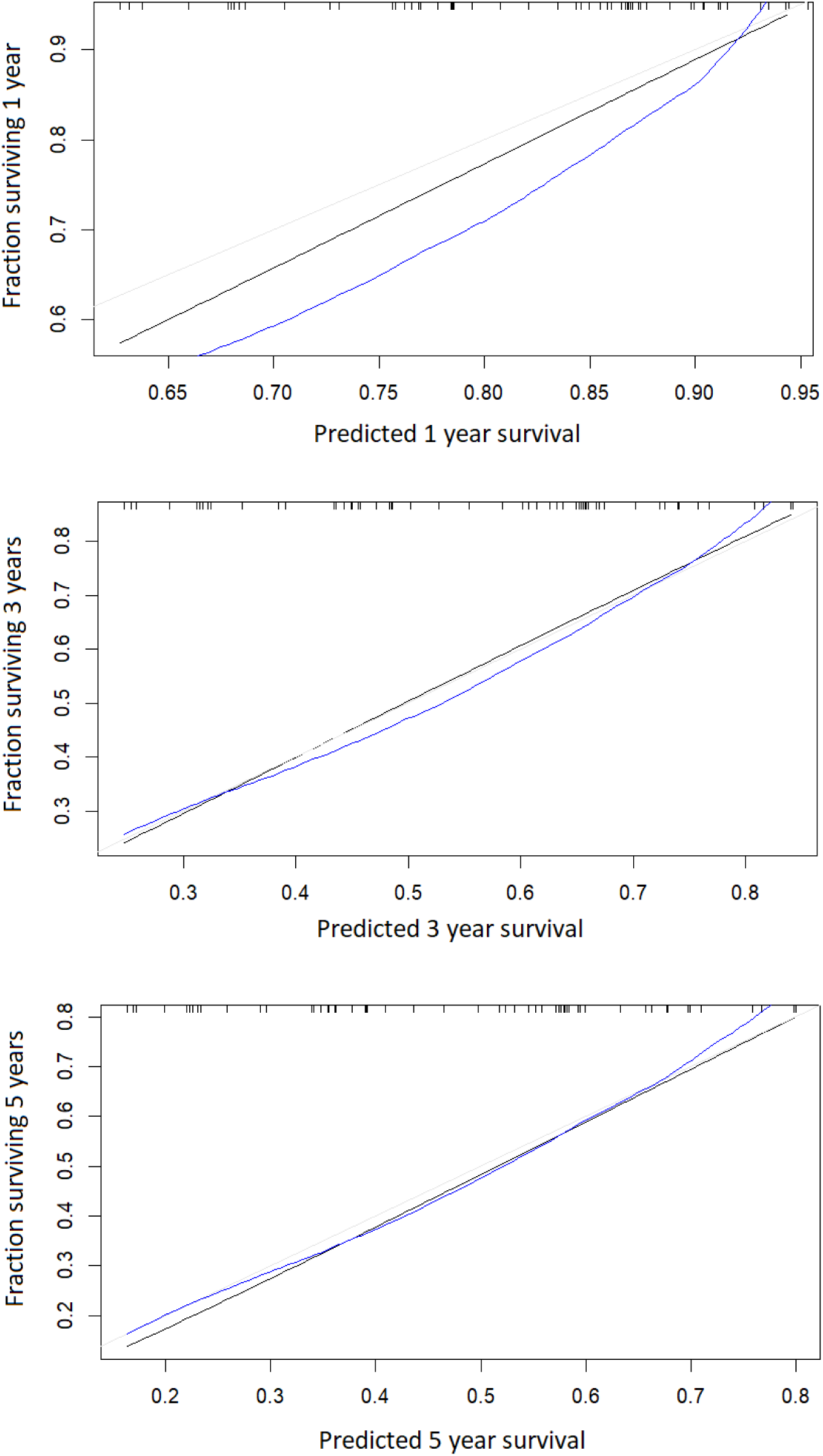
Calibration curves for CSF1 as prognostic factor for overall survival. Blue lines: with bootstrap correction; black lines: uncorrected model; grey lines: ideal fit.

**Supplemental Figure 2.**
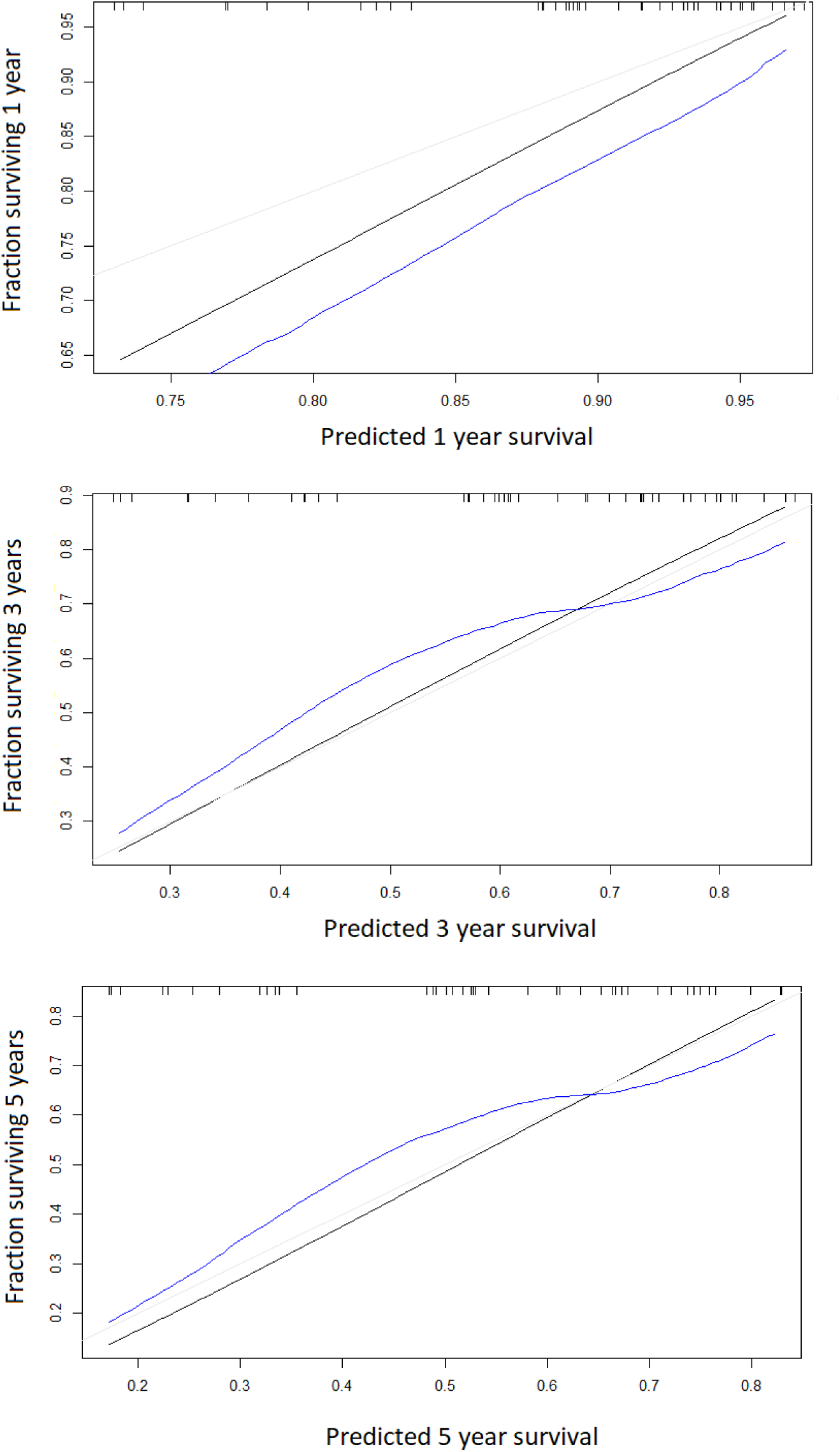
Calibration curves for CSF1 and GPS together as a prognostic model for overall survival. Blue lines: with bootstrap correction; black lines: uncorrected model; grey lines: ideal fit.

**Supplemental Figure 3.**
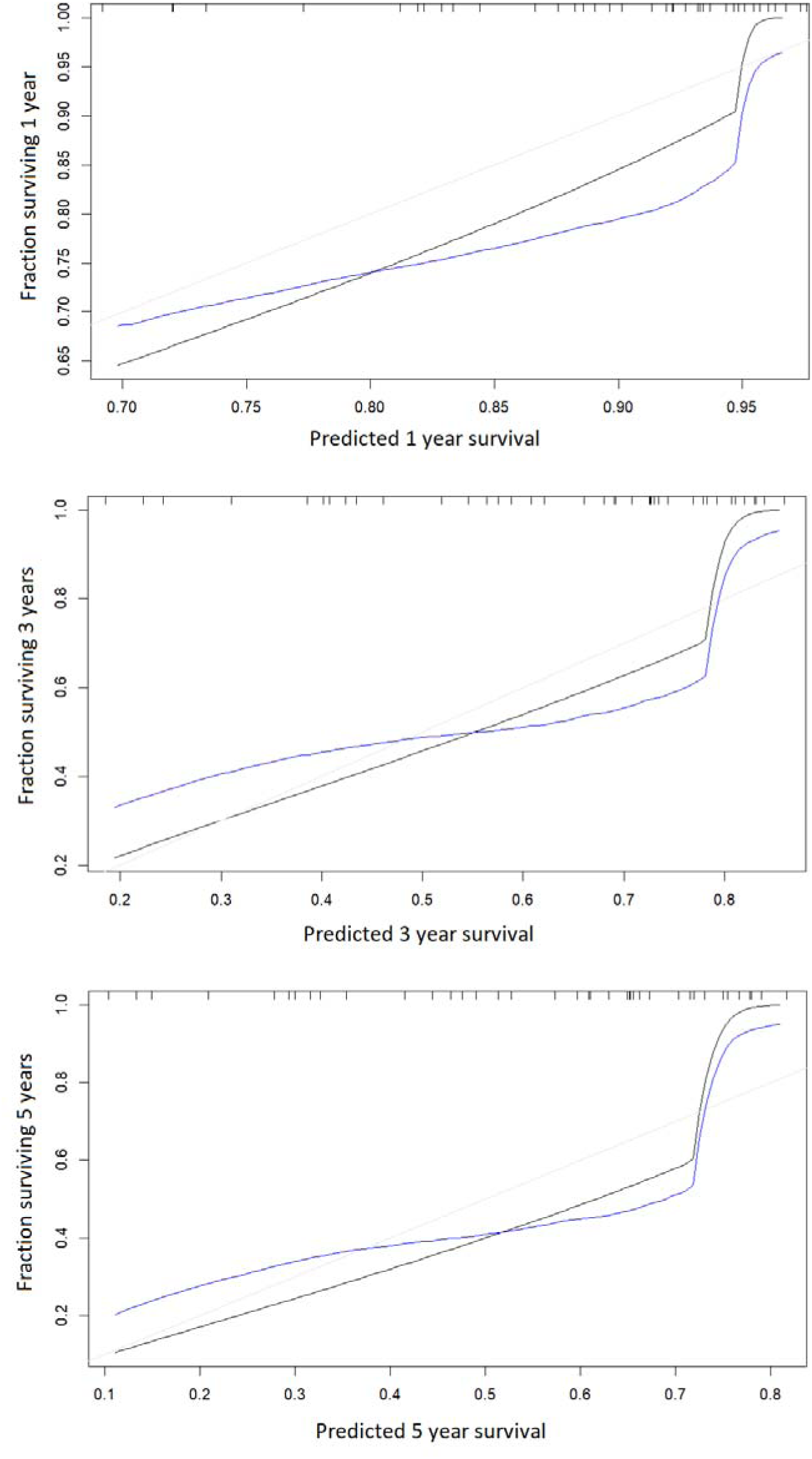
Calibration curves for CSF1, ASA and GPS together as a combined preoperative prognostic model for overall survival. Blue lines: with boot-strap correction; black lines: uncorrected model; grey lines: ideal fit

**Supplemental Figure 4.**
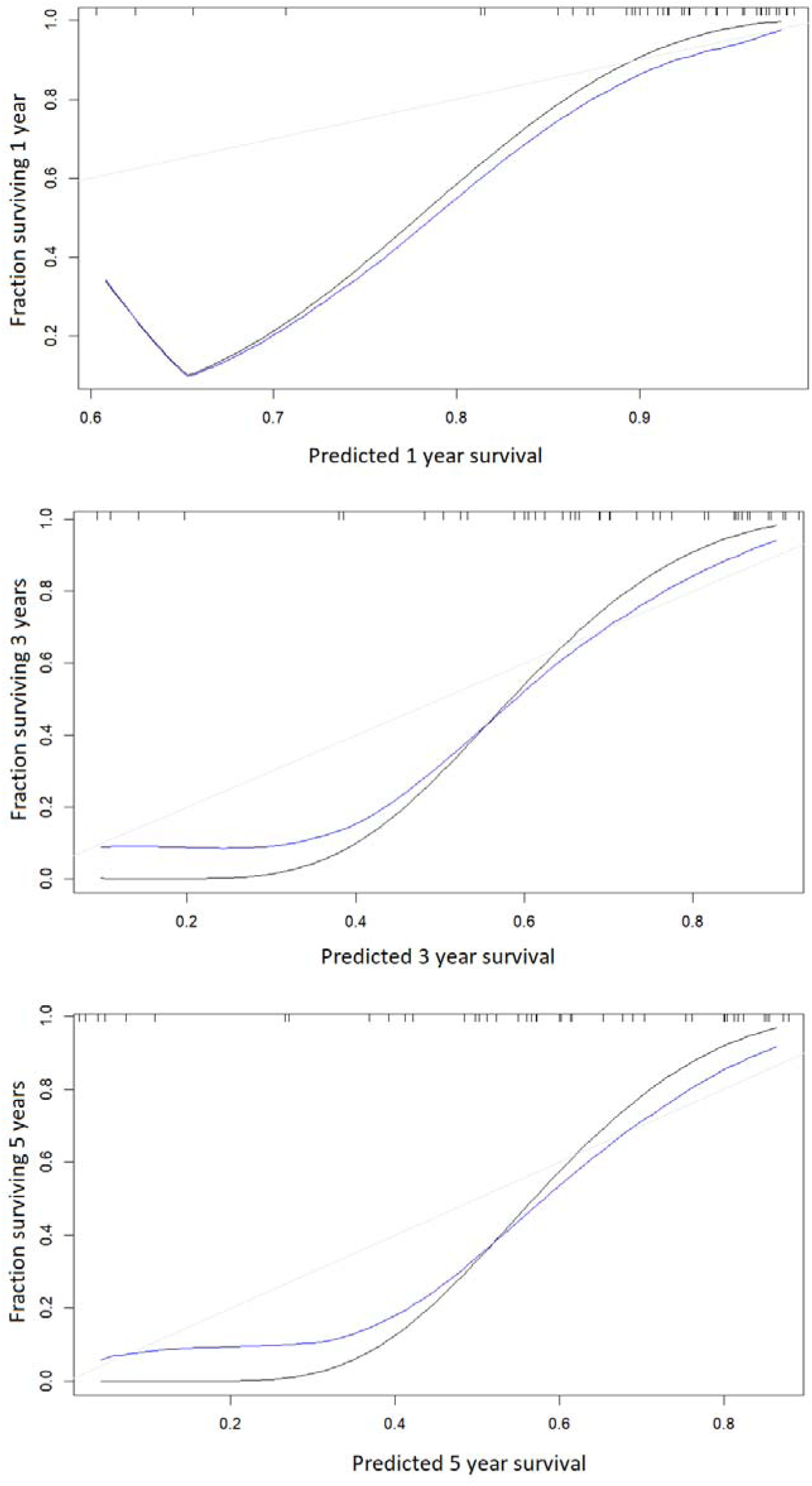
Calibration curves for CSF1, GPS, ASA and T-classification together as a total prognostic model for overall survival. Blue lines: with bootstrap correction; black lines: uncorrected model; grey lines: ideal fit.

**Supplemental Figure 5.**
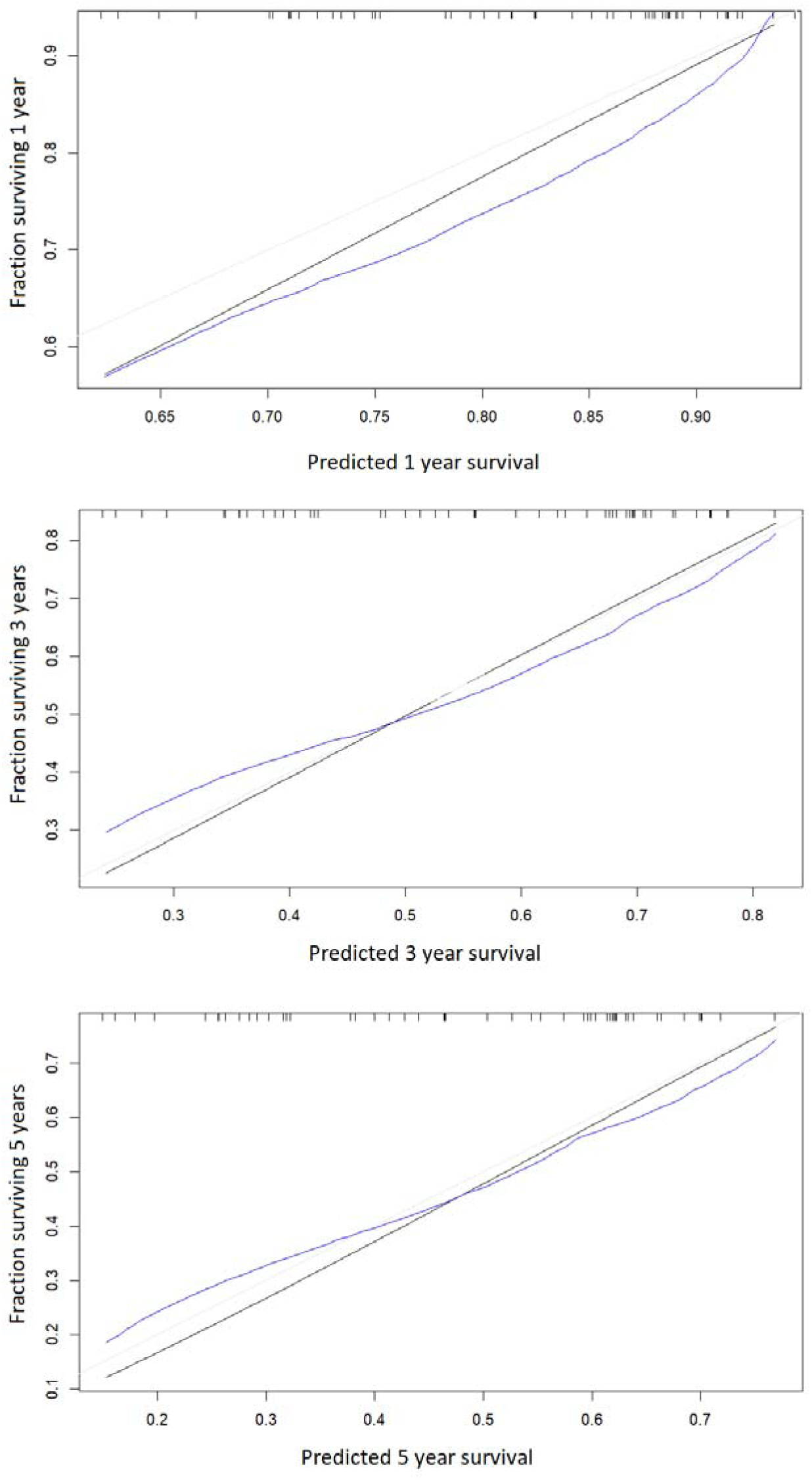
Calibration curves for CSF1 and ASA together as a total prognostic model for overall survival. Blue lines: with bootstrap correction; black lines: uncorrected model; grey lines: ideal fit.

**Supplemental Table 1.** REMARK checklist.

| Item to be reported |  | Page no. |
| --- | --- | --- |
| **INTRODUCTION** |  |  |
| 1 | State the marker examined, the study objectives, and any pre-specified hypotheses. | 5 |
| **MATERIALS AND METHODS** |  |  |
| *Patients* |  |  |
| 2 | Describe the characteristics (e.g., disease stage or co-morbidities) of the study patients, including their source and inclusion and exclusion criteria. | 6,10 and Table 1 |
| 3 | Describe treatments received and how chosen (e.g., randomized or rule-based). | 7 |
| *Specimen characteristics* |  |  |
| 4 | Describe type of biological material used (including control samples) and methods of preservation and storage. | 8-9 |
| *Assay methods* |  |  |
| 5 | Specify the assay method used and provide (or reference) a detailed protocol, including specific reagents or kits used, quality control procedures, reproducibility assessments, quantitation methods, and scoring and reporting protocols. Specify whether and how assays were performed blinded to the study endpoint. | 8-9 |
| *Study design* |  |  |
| 6 | State the method of case selection, including whether prospective or retrospective and whether stratification or matching (e.g., by stage of disease or age) was used. Specify the time period from which cases were taken, the end of the follow-up period, and the median follow-up time. | 6-7,10 |
| 7 | Precisely define all clinical endpoints examined. | 8 |
| 8 | List all candidate variables initially examined or considered for inclusion in models. | 8 |
| 9 | Give rationale for sample size; if the study was designed to detect a specified effect size, give the target power and effect size. | 6-7 |
| *Statistical analysis methods* |  |  |
| 10 | Specify all statistical methods, including details of any variable selection procedures and other model-building issues, how model assumptions were verified, and how missing data were handled. | 9 |
| 11 | Clarify how marker values were handled in the analyses; if relevant, describe methods used for cutpoint determination. | 8-9 |
| **RESULTS** |  |  |
| *Data* |  |  |
| 12 | Describe the flow of patients through the study, including the number of patients included in each stage of the analysis (a diagram may be helpful) and reasons for dropout. Specifically, both overall and for each subgroup extensively examined report the numbers of patients and the number of events. | 7, Figure 1 |
| 13 | Report distributions of basic demographic characteristics (at least age and sex), standard (disease-specific) prognostic variables, and tumor marker, including numbers of missing values. | Table 1 |
| *Analysis and presentation* |  |  |
| 14 | Show the relation of the marker to standard prognostic variables. | Table 2 |
| 15 | Present univariable analyses showing the relation between the marker and outcome, with the estimated effect (e.g., hazard ratio and survival probability). Preferably provide similar analyses for all other variables being analyzed. For the effect of a tumor marker on a time-to-event outcome, a Kaplan-Meier plot is recommended. | Table 2, Figure 2-3, Supp Table 3-4 |
| 16 | For key multivariable analyses, report estimated effects (e.g., hazard ratio) with confidence intervals for the marker and, at least for the final model, all other variables in the model. | Table 2, Supp Table 3-4 |
| 17 | Among reported results, provide estimated effects with confidence intervals from an analysis in which the marker and standard prognostic variables are included, regardless of their statistical significance. | Table 2, Supp Table 3 |
| 18 | If done, report results of further investigations, such as checking assumptions, sensitivity analyses, and internal validation. | 15-18, Supp Figure 1-5 |
| **DISCUSSION** |  |  |
| 19 | Interpret the results in the context of the pre-specified hypotheses and other relevant studies; include a discussion of limitations of the study. | 20-22 |
| 20 | Discuss implications for future research and clinical value. | 22 |

**Supplemental Table 2.** Comparison of baseline characteristics for patients with resectable (n=61) and unresectable (n=7) intrahepatic cholangiocarcinoma.

|  | Missing data | Resectable (n=61) | Missing data | Unresectable (n=7) | p-value |
| --- | --- | --- | --- | --- | --- |
| Age, md (IQR) | 0 | 69 (62-74) | 0 | 67 (61-72.5) | 0.52§ |
| Sex (male), n (%) | 0 | 34 (56) | 0 | 4 (57) | 1.00 |
| BMI, md (IQR) | 0 | 26.6 (23.7-30.0) | 0 | 25.3 (23.5- 29.8) | 0.94§ |
| ASA ≥3, n (%) | 0 | 20 (33) | 0 | 4 (57) | 0.39 |
| Diabetes, n (%) | 0 | 16 (26) | 0 | 1 (14) | 0.67<sup>#</sup> |
| PSC, n (%) | 0 | 2 (3) | 0 | 0 (0) | 1.00<sup>#</sup> |
| IBD, n (%) | 0 | 6 (10) | 0 | 0 (0) | 1.00<sup>#</sup> |
| Albumin g/L, md (IQR) | 11 | 38 (34-40) | 5 | 37.5 (35.75-39.25) | 0.96§ |
| CRP mg/L, md (IQR) | 9 | 4 (2-12) | 5 | 1.5 (1.25-1.75) | 0.09§ |
| CA19-9 kU/L, md (IQR) | 25 | 32 (13-584) | 4 | 21 (11-37) | 0.33§ |
| T ≥3, n (%) | 0 | 24 (39) | 5 | 2 (29) | 0.17<sup>#</sup> |
| N1, n (%) | 0 | 12 (20) | 4 | 3 (43) | 0.12<sup>#</sup> |
| CSF1, md (IQR) | 0 | 139 (130-154) | 0 | 128 (126-161) | 0.72§ |
| TRAIL, md (IQR) | 0 | 318 (263-373) | 0 | 298 (261-357) | 0.73§ |

**Abbreviations:** BMI, body mass index. ASA≥3, American Society of Anesthesiologists physical status classification 3-4. PSC, primary sclerosing cholangitis. IBD, inflammatory bowel disease. CRP, c-reactive protein. CA19-9, carbohydrate antigen 19-9. T≥3, tumor extension 3-4. N1, lymph node metastasis. CSF1, colony stimulating factor 1. TRAIL, TNF-related apoptosis-inducing ligand. P-values <0.05 in bold. § Mann-Whitney-Wilcoxon test. # Fisher's exact test.

**Supplemental Table 3.** Univariable Cox regression for OS and DFS according to binary variables.

|  | *Overall survival* |  |  | *Disease-free survival* |  |  |
| --- | --- | --- | --- | --- | --- | --- |
|  | HR (95% CI) | p-value | c-ind. | HR (95% CI) | p-value | c-ind. |
| GPS<br>(1-2 vs 0) | 3.16 (1.23-8.14) | **0.017** | 0.66 | 2.10 (0.91- 4.85) | 0.08 | 0.61 |
| CA19-9<br>(≥35 vs <35<br>kIE/L) | 4.84<br>(1.74- 13.51) | **0.003** | 0.69 | 4.16<br>(1.73- 9.95) | **0.001** | 0.69 |
| CSF1 quar-<br>tile 4<br>(≥158 vs<br><158 pg/mL) | 2.76 (1.05-7.30) | **0.04** | 0.65 | 2.45 (1.06- 5.66) | **0.04** | 0.60 |
| CSF1<br>(pg/mL) | 1.03 (1.01-1.05) | **0.001** | 0.71 | 1.02 (1.00- 1.04) | **0.026** | 0.64 |

**Abbreviations.** GPS, Glasgow prognostic score. CA19-9, carbohydrate antigen 19-9. CSF1, colony stimulating factor 1. P-values <0.05 in bold.

**Supplemental Table 4.**
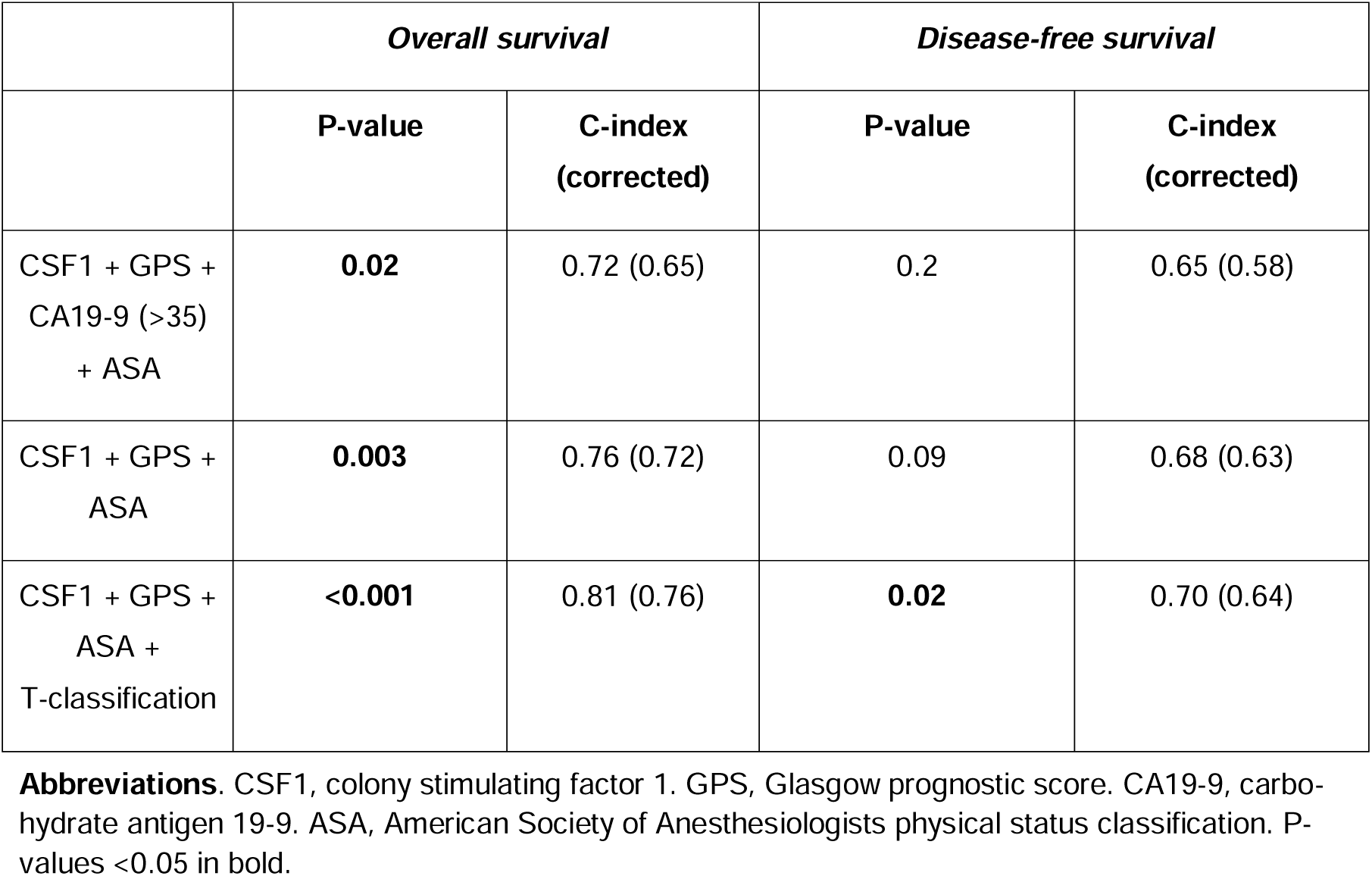
Multivariable Cox regression for OS and DFS.

|  | *Overall survival* |  | *Disease-free survival* |  |
| --- | --- | --- | --- | --- |
|  | **P-value** | **C-index**<br>**(corrected)** | **P-value** | **C-index**<br>**(corrected)** |
| CSF1 + GPS +<br>CA19-9 (>35)<br>+ ASA | **0.02** | 0.72 (0.65) | 0.2 | 0.65 (0.58) |
| CSF1 + GPS +<br>ASA | **0.003** | 0.76 (0.72) | 0.09 | 0.68 (0.63) |
| CSF1 + GPS +<br>ASA +<br>T-classification | **<0.001** | 0.81 (0.76) | **0.02** | 0.70 (0.64) |

**Abbreviations.** CSF1, colony stimulating factor 1. GPS, Glasgow prognostic score. CA19-9, carbohydrate antigen 19-9. ASA, American Society of Anesthesiologists physical status classification. P-values <0.05 in bold.

